# Non-contrast-Enhanced Functional Lung MRI to evaluate treatment response of allergic bronchopulmonary aspergillosis in patients with cystic fibrosis: a pilot study

**DOI:** 10.1101/2023.03.03.23286576

**Authors:** Ilyes Benlala, Rabea Klaar, Thomas Gaass, Julie Macey, Stéphanie Bui, Patrick Berger, François Laurent, Gael Dournes, Julien Dinkel

## Abstract

**Background:** Allergic bronchopulmonary aspergillosis (ABPA) in cystic fibrosis (CF) patients is associated with severe lung damage and requires specific therapeutic management. Repeated imaging is recommended to both diagnose and follow-up the response to treatment of ABPA in CF.

**Purpose:** To evaluate whether Fourier decomposition (FD) functional lung MRI can detect the response to treatment of ABPA in CF patients.

**Study type:** Retrospective longitudinal.

**Population:** A total of 12 CF patients.

**Field strength/sequence:** 2D balanced steady-state free precession (bSSFP) sequence with Fourier decomposition (FD) at 1.5T scanner.

**Assessment:** Ventilation weighted (V) and perfusion weighted (Q) maps were obtained after FD processing of the 2D coronal bSSFP time resolved images before and after treatment of ABPA. Defects extent was assessed on the functional maps using a visual semi quantitative score. Mean and coefficient of variation (cv) of the ventilation signal intensity (VSI) and the perfusion signal intensity (QSI) were calculated. Measurements were performed independently by two readers and averaged. The reproducibility of the measurements was also assessed. Pulmonary function tests (PFTs) were performed as markers of the airflow limitation severity.

**Statistical tests:** Comparisons of medians were assessed using paired Wilcoxon test. Reproducibility was assessed using the intraclass correlation coefficient (ICC). Correlations were assessed using Spearman test. A p value <0.05 was considered as significant.

**Results:** Defects extent on both V and Q maps showed a significant reduction after ABPA treatment (p<0.01). VSI_mean was significantly increased after treatment (p<0.01). Visual analyses reproducibility showed an ICC >0.93. ICC of the quantitative measurements was almost perfect (>0.99). VSI_cv and QSI_cv variations correlated inversely with the variation of obstructive parameters of PFTs (rho = -0.68, p=0.01).

**Data conclusion:** Non-contrast enhanced FD lung MRI appears to be able to reproducibly assess response to treatment of ABPA in CF patients and correlates with PFTs’ obstructive parameters.

## Introduction

Cystic fibrosis (CF) is one of the most frequent genetic disorders in the Caucasian population [1]. It is caused by mutations in the cystic fibrosis transmembrane conductance regulator (CFTR) gene, resulting in diverse pathologic manifestations including bronchiectasis, nasal polyposis, pancreatic insufficiency, and infertility [2]. Lung involvement (*i.e*., destruction of lung parenchyma and decline in pulmonary function) is responsible for most life-limiting complications [1]. Mucus composition and thickness in CF patients provide a favourable environment for fungal respiratory infections [3]. Indeed, *Aspergillus fumigatus* has been found in the sputum of up to 57% of patients with CF [4]. The growth of *A. fumigatus* hyphae within the bronchial lumen may be responsible for an immunoglobulin E (IgE)-mediated hypersensitivity response, known as allergic bronchopulmonary aspergillosis (ABPA), leading to bronchial inflammation and airway destruction. Its prevalence in CF patients ranges between 3 to 25%, higher than in asthmatic patients [5]. In CF patients, ABPA, which is associated with severe lung damage and requires specific therapeutic management, may be suspected due to worsening of pulmonary function and evidence of new infiltrates on chest radiographs or computed tomography (CT). Also, the findings of ABPA on chest CT include central bronchiectasis, often with mucoid impaction, centrilobular nodules, mosaic attenuation, and cavitation [6–8]. Another finding of ABPA is high-attenuation mucus plugging (HAM) on CT images, which has been considered highly specific, if not pathognomonic of the disease [9]. One of the important aspects in the management of ABPA in CF patients is repeating imaging after treatment to evaluate the effectiveness of the chosen therapeutics. However, repeating CT examinations should be limited due to the risk associated with high cumulative radiation exposure [10, 11]. Recently, lung magnetic resonance imaging (MRI) has been evaluated to assess structural alterations in CF [12, 13]. Indeed, thanks to recent breakthroughs, including ultra-short echo time (UTE) MRI sequences, lung morphological modifications in CF assessed using UTE-MRI have been found similar to that of CT [12, 14]. Furthermore, inverted mucus impaction signal on MRI (IMIS), which is characterized by a high signal intensity on T1-weighted images and low signal intensity on T2-weighted images, can be considered as the counterpart of HAM and has been found 100% specific to detect ABPA in CF patients [15]. Moreover, lung MRI can evaluate pulmonary functional changes in a regional fashion [16]. Non-contrast techniques of pulmonary ventilation-perfusion assessment in CF patients (*e.g*., Fourier decomposition (FD) MRI) have been reported [17, 18]. They have been validated vs. photon emission computed tomography (SPECT), dynamic contrast-enhanced MRI and hyperpolarized 3He, considered as the gold standard in assessing lung function using imaging [19]. Nonetheless, longitudinal evaluations before and after treatment of ABPA in CF patients using morphological and functional lung MRI are lacking. We hypothesised that FD-MRI can detect response to treatment in CF patients with ABPA. Thus, the main objective of this study was to evaluate the response to treatment of ABPA in CF patients based on functional changes. As secondary objectives, we evaluated morphological changes before and after treatment of ABPA, we assessed correlations between both morphological and functional alterations and pulmonary function tests (PFTs) parameters in addition to the structure-function relationships in CF patients with ABPA using UTE-MRI and FD-MRI.

## Material and methods

### Study population

This retrospective longitudinal pilot study was performed in a single center and approved by the local ethics committee while written consent was waved. All consecutive CF patients referred to our institution, a tertiary CF center, were screened between August 2018 and September 2021. Inclusion criteria were: diagnosis of CF proven by sweat chloride and/or genetic testing, age older than 6 years, diagnosis of ABPA established by multidisciplinary sessions involving pediatricians, pneumologists, physicians, immunologists, physiologists, and mycologists with full knowledge of the patient’s medical history and according to the CFFC criteria [20]. All included patients underwent lung MRI with UTE and FD sequences before and after treatment of ABPA. Median interval between the two examinations was 5 months (minimum of 3 months and maximum of 9 months). In addition, total IgE and anti–*A fumigatus* specific IgE levels were measured before and after treatment as well as pulmonary function tests within one week from the lung MRI examination. Clinical and radiological improvement and/or a drop of at least 25% of total IgE were considered as positive response to ABPA treatments [21].

### Pulmonary Function Tests

PFTs were assessed using body plethysmography (BodyBox, Medisoft. Leeds, UK). Reference values were based on American Thoracic Society and European Respiratory Society guidelines [22, 23]. Measurements of forced expiratory volume in 1 second (FEV1), forced volume capacity (FVC), FEV1/FVC and forced expiratory flow at 25%-75% (FEF_25-75_) were measured and expressed as percentage of predicted values.

### MRI examinations

MRI examinations were completed on a Siemens Aera (Siemens, Erlangen, Germany) at 1.5 Tesla. Images of the stack of spirals spoiled ultra-short gradient echo sequence (3D-UTE) were acquired in the supine position using the following parameters: TR/TE/flip angle=4.3ms/0.05ms/5° and a voxel size of 1mm^3^. Respiratory synchronization at end normal expiration was allowed by an automated self-navigated respiratory gating. Scan duration was between 6 and 8 minutes. Time resolved images of the 2D balanced steady-state free precession (bSSFP) sequence were acquired at two coronal positions: at the level of the carina and posteriorly at the level of the descendant aorta. The main pulse sequence parameters were: TR/TE/flip angle=1.74ms/0.71ms/47°, pixel size of (1.2×1.2 mm)^2^ and slice thickness = 12 mm. The series consisted of 200 images. Scan duration was around 1 minute per slice. In addition, for IMIS identification, T2-weighted radial fast spin-echo (RFSE) sequence was performed using the following parameters: TR/TEs/flip angle=2350ms/20ms-150ms/145°; pixel size of (1.6 × 1.6) mm^2^ and slice thickness = 1.6mm. T1-weighted VIBE sequence was also performed using the following parameters: TR/TEs/flip angle=3.37ms/1.28ms/8°; pixel size of (1.2 × 1.2) mm^2^ and slice thickness = 3mm.

### Fourier decomposition (FD) image Processing Workflow

The image processing workflow was fully implemented in Python (version 3.9). After discarding the first 20 images, where the steady state condition was not fulfilled [17], the acquired image series were first elastically registered to a reference image in mid-position between full inspiration and full expiration determined automatically using average lung signal variation. The deformable registration was applied using the EVolution algorithm [24], which is based on a similarity term that favours edge alignment and on a diffeomorphic transformation that ensures the preservation of the image topology. Based on this reference image, a manual segmentation of the lung was performed as well as a region of interest (ROI) drawn within the aorta by two radiologists. The sampling times were used to calculate the fast Fourier Transform (FFT) per pixel on the segmented lung. The ventilation weighted (V) and perfusion weighted (Q) maps were then generated by taking the maximum magnitude of the corresponding peak in the Fourier spectrum using the lung ROI for ventilation and the aorta ROI for perfusion.

Finally, the signal within the V-map and the Q-map was normalized using the mean signal of the aorta ROI of the reference image for each examination.

A muscular ROI was also drawn on the reference image of every examination to check if the overall signal varied between the two examinations before and after treatment.

### Functional analysis

#### Qualitative evaluation

Defect extents on V and Q maps were assessed using a semi quantitative score (0= absence/negligible, 1=<50%, 2=>50%) regarding the right/left lungs, superior, median or inferior regions for each slice. The score was then averaged between the two acquired slices and ranged between 0 and 12.

#### Quantitative evaluation

Mean and coefficient of variation (cv) of the signal intensity of both ventilation and perfusion maps were calculated (*i.e*., VSI_mean, VSI_cv, QSI_mean and QSI_cv).

### Morphological analysis

A widely used structural alterations scoring system in CF patients (Bhalla score) was performed [25]. Detection of IMIS was also part of the evaluation [15].

Two radiologists with 5 and 10 years in thoracic imaging performed morphological and functional evaluations. All the measurements performed in this study were averaged between the two readers.

### Structure-function evaluation

For the structure-function relationships evaluation, a matched 3D UTE-MRI coronal slice was assessed in terms of structural changes (*i.e*., bronchiectasis without mucus plugging, bronchiectasis filled with mucus plugging and consolidation) alongside with V and Q maps. Regions were then classified into defect region or no defect region and the predominant structural alteration identified. A consensus reading was carried out for these evaluations and scrolling through the adjacent slices of the 3D UTE images was allowed for confident identification of structural changes.

### Statistical analysis

Statistical analyses were performed using MedCalc software (Version 20.216). Distribution normality was assessed using Shapiro-wilk test. Data were expressed as medians with [minimum to maximum range] for continuous variables and absolute numbers for categorical variables. Comparisons of paired medians were performed using the Wilcoxon signed-rank test and paired percentages using the McNemar test; correlations were assessed using the Spearman test. A p-value<0.05 was considered statistically significant. Reproducibility was assessed using intraclass correlation coefficients (ICCs), with mixed model analysis and absolute agreement option. ICC values were classified as null (=0), slight (>0 and <0.20), fair ≥0.20 and <0.40), moderate (≥0.40 and <0.60), good (≥0.60 and <0.80), very good (≥0.80 and <0.95) and almost perfect (≥0.95) [26]. Structure-function relationships were evaluated using Chi2 tests.

## Results

### Study population

Among 109 CF patients referred to our institution between 2018 and 2021, 29 patients showed an exacerbation attributed to ABPA according to the multidisciplinary care meeting and CFFC criteria. Seventeen patients were not included (nine did not undergo lung MRI before treatment and eight did not have lung MRI after treatment within a maximum interval of one week from the IgE measurements). Twelve patients were then included in this study after they have had lung MRI, total IgE and specific IgE measurements and PFTs before and after treatment of ABPA (Figure 1). All included patients showed a positive response to specific treatment (*i.e*., oral corticosteroids and itraconazole) either with a drop of total IgE or clinical and radiological improvement or both. Patients’ characteristics are summarized in Table 1. Median age of the included patients before treatment was 14 years with 7 males and 5 females. The medians of total IgE and specific IgE before treatment were 667.5 UI/ml and 10.5 UI/ml respectively with a significant reduction at 467.5 UI/ml and 8.36 UI/ml respectively after treatment (p<0.01). FEV1%p showed a significant improvement after ABPA treatment with a median FEV1%p increasing from 61% to 67% (p<0.01).

**Figure 1:**
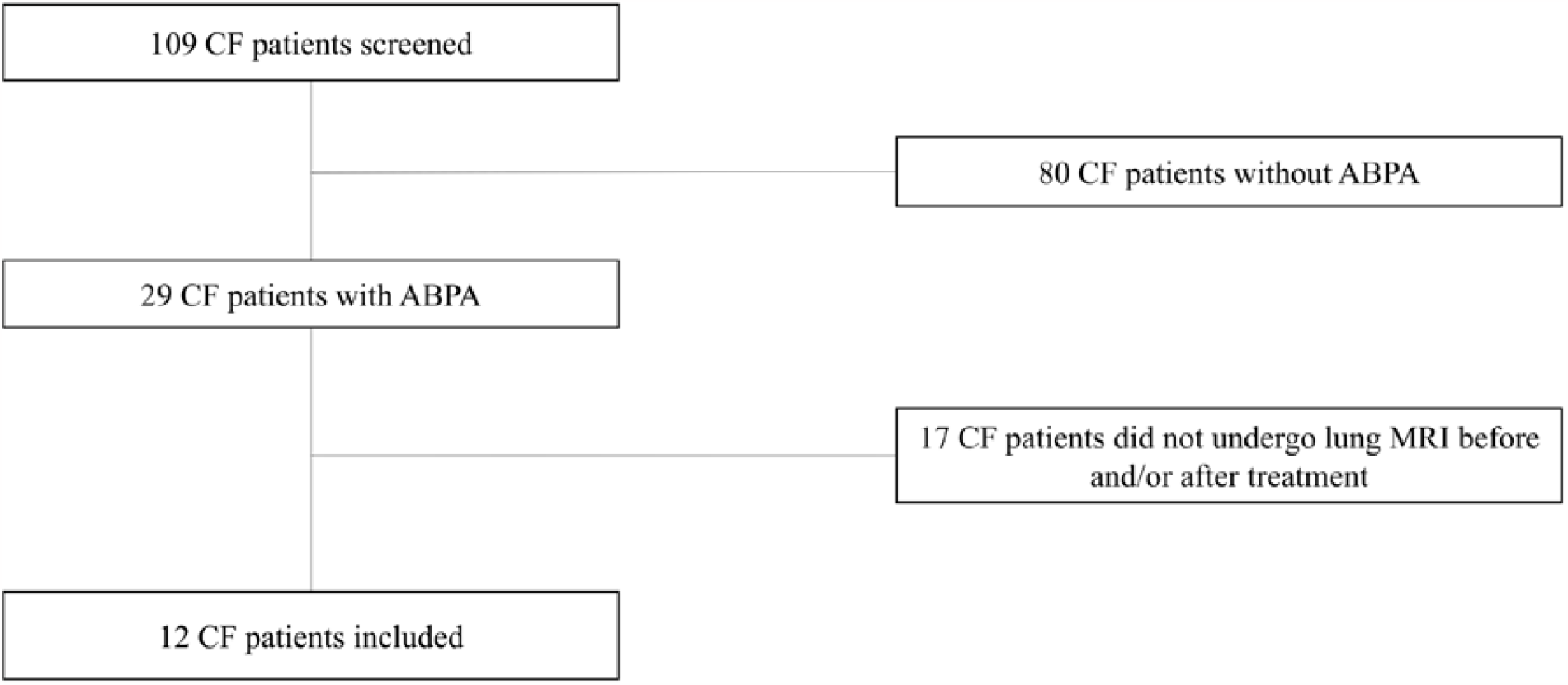
Flowchart of the study. CF= cystic fibrosis; ABPA= allergic bronchopulmonary aspergillosis.

**Table 1.** Clinical and functional characteristics of study participants.

|  |  | **Before Treatment** | **After Treatment** | **p-value** |
| --- | --- | --- | --- | --- |
|  |  | (n=12) | (n=12) |  |
| **Age** | Years | 14 [6.5-32] | 14 [7-33] | <0.01 |
| **Sex ratio** | Male/Female | 7 / 5 | 7 / 5 | 0.77 |
| **BMI** | kg.m<sup>-2</sup> | 18.5 [13-22] | 19 [14-23] | 0.95 |
| **Mutation**<br>**ΔF508** | Homozygous<br>(yes/no) | 7 / 5 | 7 / 5 | 0.77 |
| **Total IgE** | UI/ml | 667.5 [221-3529] | 467.5 [26-1917] | <0.01 |
| **Anti-*A. fumigatus***<br>**Specific IgE** | UI/ml | 10.5 [2.15-32.3] | 8.36 [0-25.3] | <0.01 |
| **PFT** | FEV<sub>1</sub> (%p) | 61 [20-106] | 67 [22-126] | <0.01 |
|  | FVC (%p) | 78 [30-107] | 93 [32-131] | <0.01 |
|  | FEV<sub>1</sub>/FVC | 74 [55-100] | 71 [51-103] | 0.84 |
|  | FEF<sub>25-75</sub> (%p) | 28 [8-95] | 32.5 [9-104] | 0.15 |

Note: Data are median with [minimum to maximum range] for continuous variables and absolute number for categorical variables. Legends: ABPA= allergic bronchopulmonary aspergillosis; BMI=body mass index; IgE= immunoglobulin E; FEV<sub>1</sub>=forced expiratory volume in 1 second; FVC=forced volume capacity; FEF<sub>25-75</sub>=forced expiratory flow at 25%-75%; %p=percentage of predicted value

### Comparison of functional FD-MRI ventilation and perfusion before and after ABPA treatment

The extent of the ventilation and perfusion defects decreased significantly after ABPA treatment (p<0.01) reflecting a significant improvement of the lung function evaluated using FD-MRI (Table 2, Figure 2 and Figure 3).

**Table 2.** Imaging characteristics of study participants.

|  |  | **Before Treatment** | **After Treatment** | **p-value** |
| --- | --- | --- | --- | --- |
|  |  | (n=12) | (n=12) |  |
| **Visual Imaging** |  |  |  |  |
| *Functional evaluation* | V defect score | 4.125 [3-10.75] | 2 [1-7.25] | <0.01 |
|  | Q defect score | 5 [2.5-10.75] | 2.75 [0-7] | <0.01 |
| *Morphological evaluation* | MR-Bhalla score | 12.75 [9-21] | 15.5 [9.5-21] | 0.02 |
|  | IMIS (present/absent) | 6/ 6 | 0/ 12 | 0.03 |
| **Quantitative Imaging** |  |  |  |  |
|  | VSI_mean (au) | 167 [100-532] | 280 [112-535] | <0.01 |
|  | VSI_cv | 1.18 [0.96-2.09] | 1.16 [0.62-1.76] | 0.33 |
|  | QSI_mean (au) | 112 [53-298] | 122 [46-282] | 0.85 |
|  | QSI_cv | 1.32 [0.99-1.97] | 1.30 [0.82-1.92] | 0.62 |

Note: Data are median with [minimum to maximum range] for continuous variables and absolute number for categorical variables. Legends: MR= magnetic resonance; IMIS= inverted mucus signal intensity; V= ventilation; Q= perfusion; VSI= ventilation signal intensity; QSI= perfusion signal intensity; au= arbitrary unit; cv= coefficient of variation

**Figure 2:**
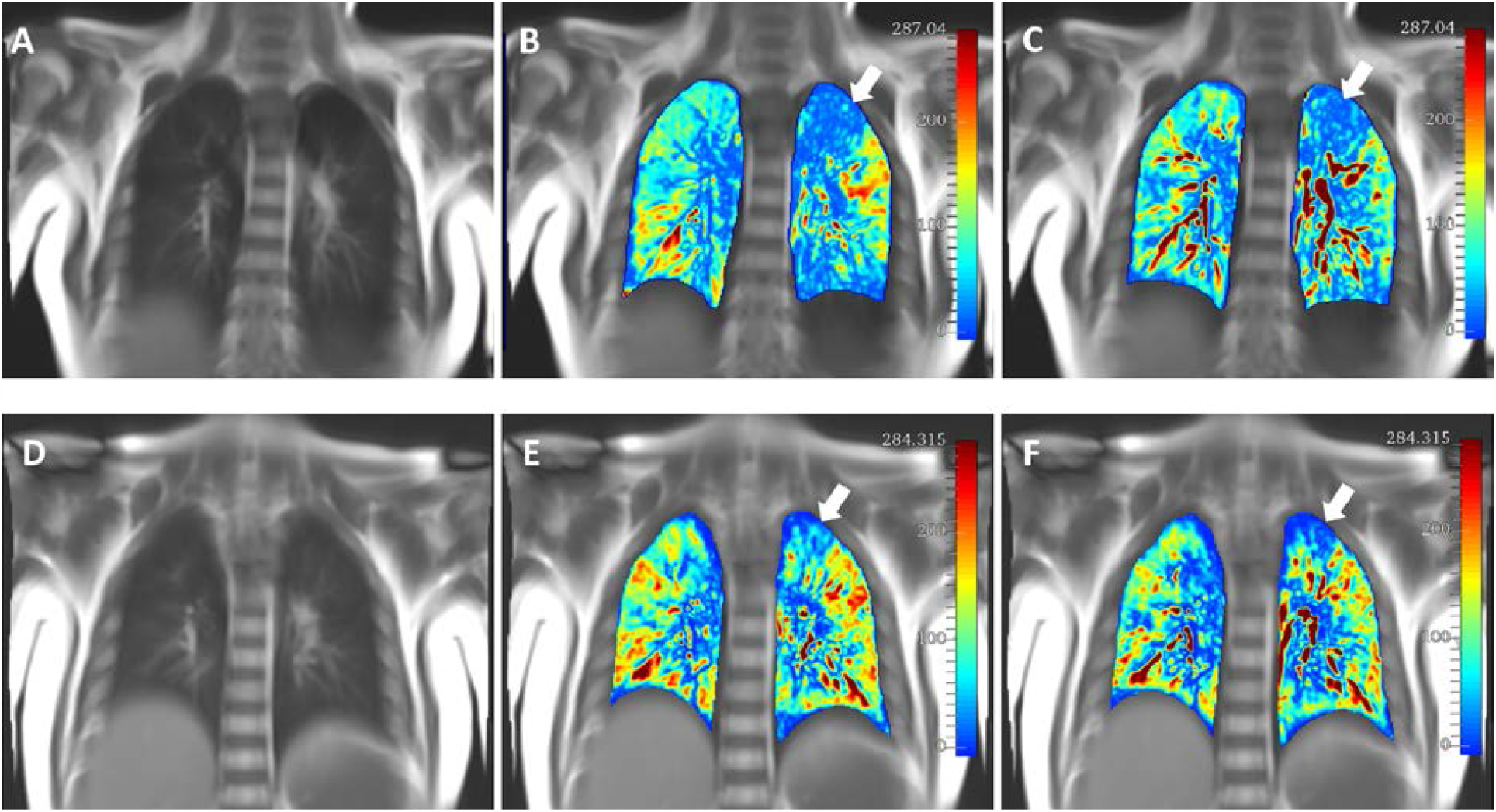
Coronal 2D balanced steady-state free precession image of the lungs before (A) and after (D) ABPA treatment. Note the matched defect of ventilation (B) and perfusion (C) before specific treatment (white arrows) with the decrease of the extent of the defects after treatment on both ventilation map (E) and perfusion map (F).

**Figure 3:**
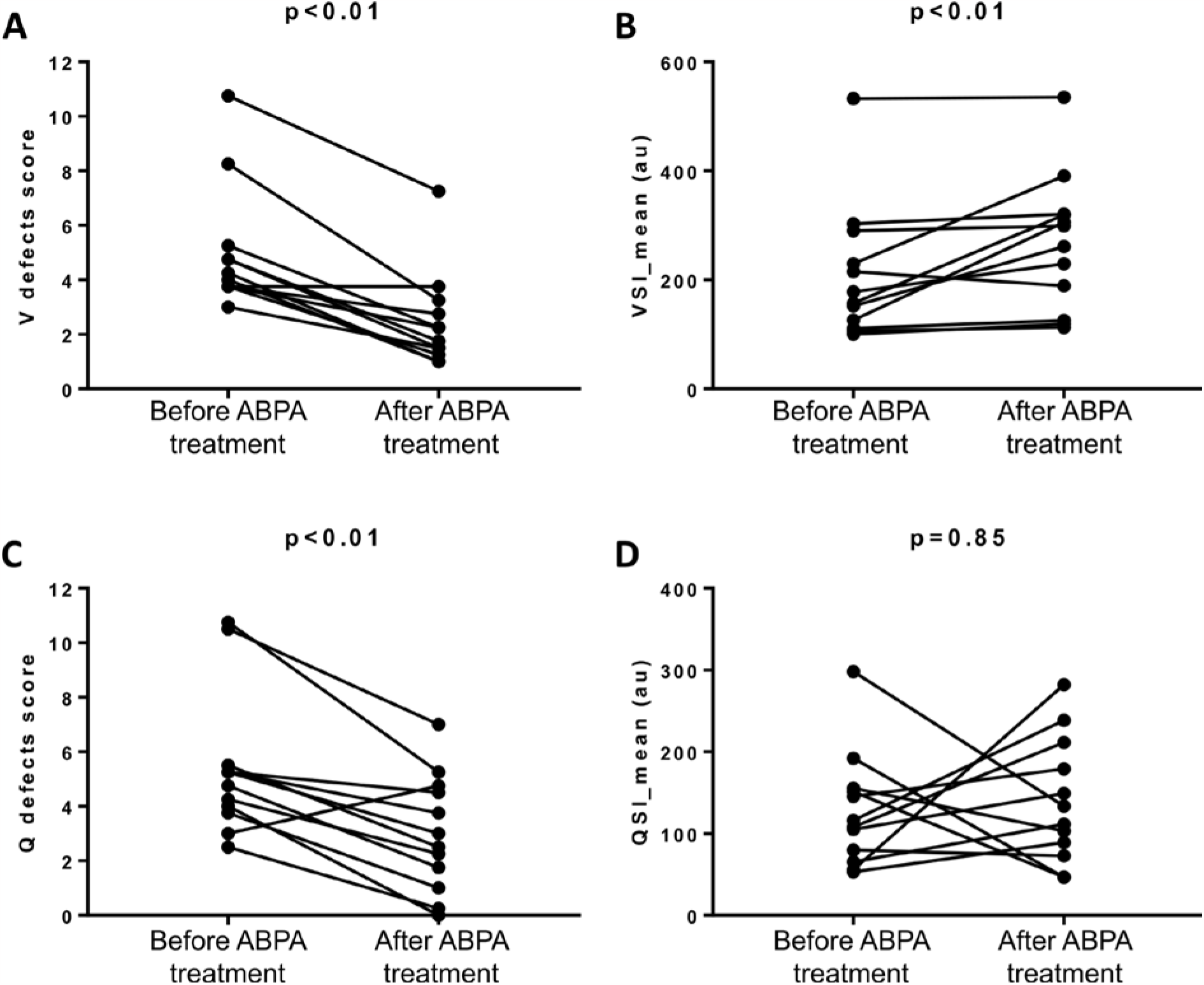
Paired comparisons of the visual and quantitative functional FD-lung MRI before and after ABPA treatment. ABPA= allergic bronchopulmonary aspergillosis ; V= ventilation; Q= perfusion; VSI= ventilation signal intensity; QSI= perfusion signal intgensity ; au= arbitrary unit.

Using the fully automated quantification of lung ventilation, the median of VSI_mean was significantly increased after ABPA treatment (p<0.01) (Table 2, Figure 3) reflecting a global improvement of the lung ventilation whereas QSI_mean showed only a trend for increase after treatment (Table 2, Figure 3). Regarding the heterogeneity of V map and Q map, the VSI_cv and QSI_cv did not demonstrate a significant improvement after ABPA treatment (p=0.33 and p=0.62, respectively) (Table 2).

No statistically significant difference was found between the mean and the standard deviation of the signal in the muscular ROIs before and after treatment (p=0.42) (Table E1).

### Comparison of morphological Bhalla score before and after ABPA treatment

The median of MR-Bhalla score was significantly increased after ABPA treatment (p=0.02) reflecting a decrease of structural alterations’ severity assessed using 3D UTE morphological MRI. In addition, 6 patients showed an IMIS on lung MRI before treatment that all disappeared on the follow-up MRI (p=0.03) (Table 2).

### Correlations with the pulmonary function tests

Before treatment, no significant correlation was found between PFT’s parameters and lung MRI functional or morphological measurements. After treatment, ventilation defects extent assessed visually was inversely correlated to PFT’s obstructive parameters (Table E2). Using quantitative imaging, QSI_mean was significantly correlated to FEV1%p and QSI_cv showed a significant inverse correlation with FEF25-75%p (Table E2). The MR-Bhalla score also showed a significant correlation with PFTs’ obstructive parameters after treatment (Table E2).

However, only quantitative functional imaging parameters (*i.e*., ΔVSI_cv and ΔQSI_cv) showed significant correlations with the variation of PFTs’ obstructive parameters (ΔFEV1%p and ΔFEF25-75%p) (Figure 4, Table E3).

**Figure 4:**
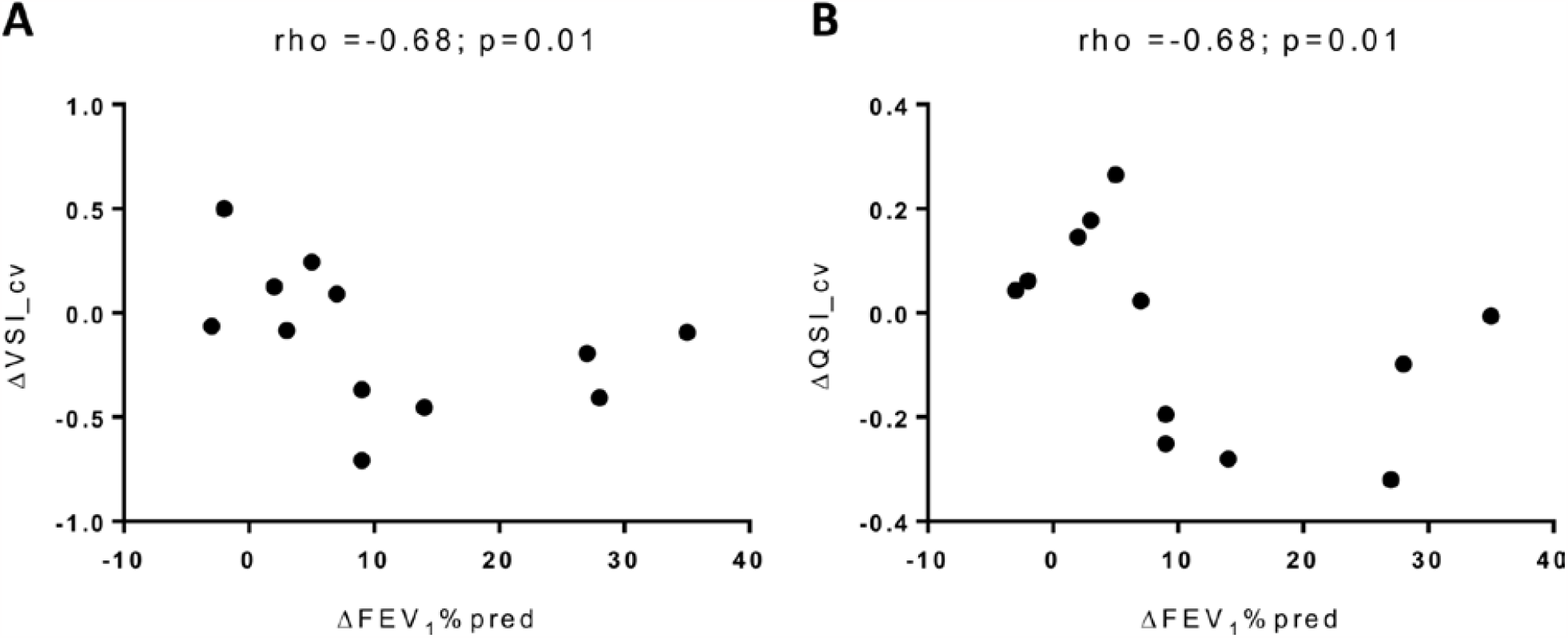
Spearman correlations between the variation of FEV1%p and the variation of both ventilation and perfusion coefficent of variation (VSI_cv and QSI_cv).

### Assessment of reproducibility

Very good inter-observer reproducibility were obtained for functional and morphological visual scores (ICC>0.93). Reproducibility of quantitative measurements were almost perfect, with ICCs >0.99 (Table 3).

**Table 3.**
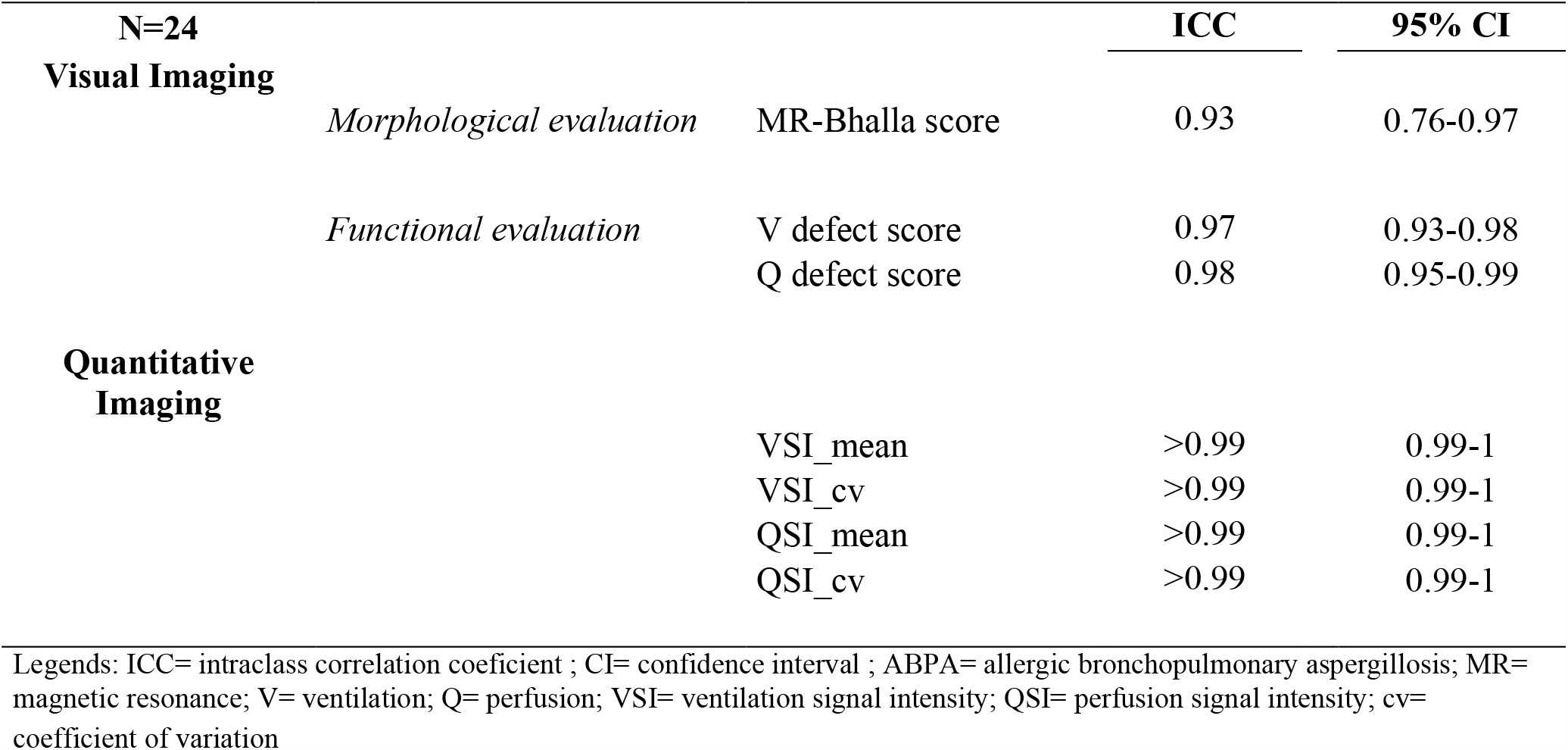
Interobserver reproducibility of visual and quantitative analyses.

### Structure-function relationships

A total of 576 regions were evaluated. Three hundred and twenty-two (322) regions were found without any defect and 254 with identified defects (130 with Q defects and 124 with V defects). Regarding the defect extent, 115 defects were found with less than 50% extent and 139 with more than 50% extent. According to structural alterations evaluated in this study (Table 4), the presence of bronchiectasis without mucus plugging were found in 13 regions without any defects, and no bronchiectasis without mucus plugging was found in region with V or Q defects. Bronchiectasis with mucus plugging were found in 39 regions without any defect and 232 regions with V or Q defects (Figure 5). Consolidations were found only in regions with V or Q defects (18 regions). Only 4 regions showed functional defects (1 with V defects and 3 with Q defects) without any visible structural alteration. Presence of structural alterations was not different between regions with V defects and regions with Q defects (p=0.06). The presence of functional defects (V or Q) was different between regions with structural alterations and regions with no visible structural alterations (p<0.001). In addition, the presence of functional defects (V or Q) was different according to the structural alterations assessed (p<0.001). Only 6 regions showed a mismatch between V and Q defects (p=0.33).

**Table 4.** Structure-function evaluation in 576 lung regions.

|  | **No defects** | **V defects** |  | **Q defects** |  |
| --- | --- | --- | --- | --- | --- |
|  |  | <50% | >50% | <50% | >50% |
| **Bronchiectasis without MP** | 13 | 0 | 0 | 0 | 0 |
| **Bronchiectasis with MP** | 39 | 49 | 65 | 52 | 66 |
| **Consolidation** | 0 | 5 | 4 | 5 | 4 |
| **No visible structural alteration** | 270 | 1 | 0 | 3 | 0 |

Data are absolute numbers. Legends: V= ventilation, Q= perfusion, MP= mucus plugging

**Figure 5:**
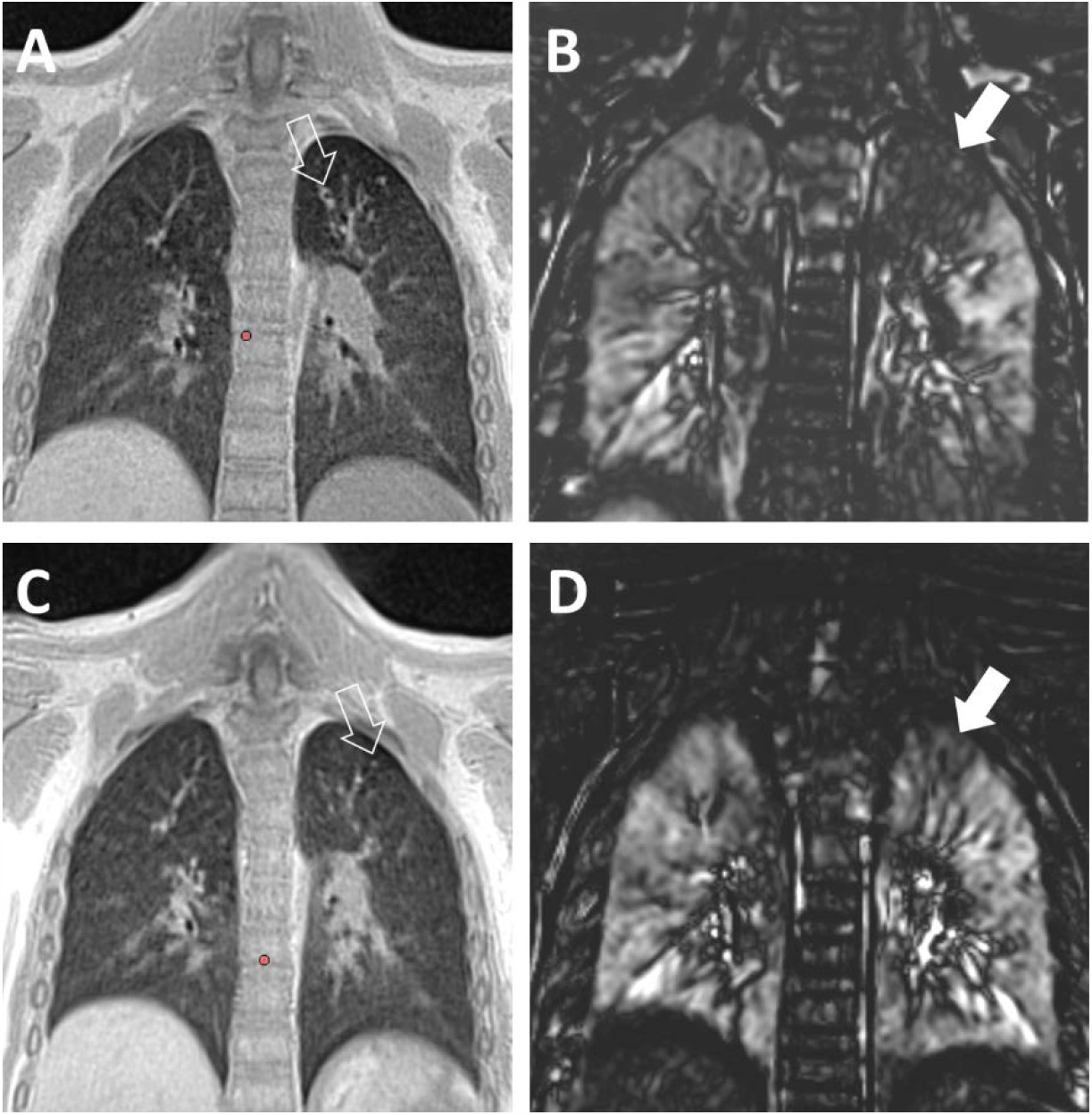
Matched coronal slice of the lung UTE-MRI image and FD ventilation map before (A, B) and after (C, D) ABPA treatment. Note the ventilation defect indicated by the solid arrows before (B) and after (D) treatment and the structural alterations (hollow arrows) seen on the UTE-MRI images showing mucus plugging (A) decreasing after treatment (C).

Moreover, no correlation was found between the MR-Bhalla morphological score and both visual and quantitative evaluations of ventilation and perfusion (p>0.30) (Table E4).

## Discussion

We demonstrated in this study that lung functional FD-MRI is able to assess ventilation and perfusion defects in CF patients with ABPA. We also showed that automatic quantification of lung V and Q is feasible in CF patients with ABPA using FD-MRI. Our findings showed that the extent of V and Q defects decreased after positive response to treatment of ABPA in CF patients. Moreover, we found an increase of VSI_mean after treatment. Our visual and automatic methods showed a very good to almost perfect inter-reader reproducibility. Interestingly, only the variations of VSI_cv and QSI_cv, reflecting a more homogenous V and Q maps, were found to correlate with the variation of FEV1%p after ABPA treatment. Finally, we showed that functional defects are associated with structural alterations, especially with mucus plugging and consolidations.

To the best of our knowledge, this is the first study aiming to monitor treatment response of ABPA using non-contrast enhanced functional lung MRI. However, regarding morphological lung MRI, our findings are in line with the already reported results showing a complete resolution of IMIS in the follow-up [15]. Moreover, a recent study [27] showed that free breathing phase resolved functional lung MRI imaging can detect improvement following exacerbation treatments using antibiotics out of the context of ABPA. Furthermore, recent studies demonstrated the benefit of using other MRI techniques (*i.e*., diffusion weighted imaging and T2 weighted imaging) in monitoring CF exacerbations [28, 29].

In this study, functional defects were found to be associated with ABPA exacerbation and showed a significant decrease after specific treatment. Indeed, CF patients with ABPA are prone to more severe exacerbations and require a specific treatment with repeated imaging. However, iterative radiation exposure raises concerns in young CF patients with a need of a radiation-free alternative. Moreover, FD-MRI is performed without any contrast agent administration, representing a safer technique regarding long-term effects of MRI contrast deposition [30].

In addition, automatic quantification of VSI and QSI showed an improvement of global respiratory function after specific ABPA treatment. We used a simple normalization technique to account for signal intensity bias in longitudinal examinations and we also checked that the signal in a muscular ROI was not different between the two examinations. Interestingly, the variations of VSI_cv and QSI_cv, representing the homogeneity of V and Q maps, were correlated to the variation of FEV1%p. Thus, the decrease of the number and the extent of functional defects is associated with more homogenous maps and therefore PFTs obstructive parameters were found improved. Indeed, insights into local and regional respiratory function allowed by the FD-MRI might bring complementary information to the global evaluation of the respiratory function using PFTs in addition to the morphological information allowed by using 3D UTE-MRI.

Morphological MR-Bhalla score was also found improved in the follow-up, which is in line with the already published data of structural changes in the follow-up of exacerbations in CF patients [31–34].

In our study, we found that functional defects are associated with structural lung alterations without any distinction between V defects and Q defects. However, bronchiectasis without mucus plugging were not associated with functional defects assessed by using FD-MRI. Indeed, airway obstruction is more likely responsible for matched ventilation-perfusion defects. However, since we did not assess the relationship between the severity of the bronchiectasis and functional defects, our findings need to be interpreted with caution.

This study has some limitations. This is a retrospective pilot study in a small number of patients. Indeed, we paid a careful attention to include only patients with high confidence diagnosis of ABPA and with IgE serological measurements performed before and after treatment close to the MRI examinations. However, large prospective cohorts to validate our findings are needed. We did not correlate FD-MRI findings to dynamic contrast enhanced (DCE) MRI or to hyperpolarized gas MRI. However, functional FD-MRI was already validated using these techniques [19, 35]. Our visual evaluation of functional defects showed a high reproducibility. Nevertheless, it could be worth using an automatic relative quantification of defects based on threshold techniques [36]. Finally, a further comparison of different non-rigid image registration algorithms can provide information about the robustness of these findings regarding preprocessing techniques.

To conclude, functional changes in CF patients with ABPA can be reproducibly assessed using FD lung MRI and correlate with pulmonary function tests variation after specific treatment of ABPA. Therefore, non-contrast enhanced functional FD lung MRI might be used to monitor treatment of ABPA in CF patients.

## Data Availability

All data produced in the present study are available upon reasonable request to the authors

## Acknowledgments

This study was conducted in the framework of the University of Bordeaux’s IdEx “Investments for the Future” program RRI “IMPACT” which received financial support from the French government. The authors thank Dr Beaudouin Denis DeSenneville for technical support.

## Supplementary Tables

**Table E1.** Comparison of muscular ROI mean signal intensity and standard deviation before and after ABPA treatment.

|  | **Before Treatment** | **After Treatment** | **p-value** |
| --- | --- | --- | --- |
|  | (n=12) | (n=12) |  |
| ROI's mean (au) | 238 [84-293] | 245 [135-325] | 0.42 |
| ROI's standard deviation (au) | 9.5 [4.3-22.3] | 8.5 [2.5-22.7] | 0.85 |

Note: Data are median with [minimum to maximum range] for continuous variables. Legends: ABPA= allergic bronchopulmonary aspergillosis; ROI= region of interest, au= arbitrary units.

**Table E2.** Correlation between MRI metrics with pulmonary function test before and after treatment of ABPA.

|  |  |  |  | FEV1%p |  | FEV1/FVC |  | FEF25-75%p |  |
| --- | --- | --- | --- | --- | --- | --- | --- | --- | --- |
| | | | | $\rho$ | p-value | $\rho$ | p-value | $\rho$ | p-value |
| Visual Imaging | *Morphological evaluation* | MR-Bhalla score | Before | 0.24 | 0.45 | 0.07 | 0.83 | 0.19 | 0.54 |
|  |  |  | After | 0.64 | **0.02** | 0.24 | 0.45 | 0.64 | **0.02** |
|  | *Functional evaluation* | V defect score | Before | -0.09 | 0.76 | -0.46 | 0.13 | 0.02 | 0.94 |
|  |  |  | After | -0.69 | **0.01** | -0.68 | **0.01** | -0.73 | **0.006** |
|  |  | Q defect score | Before | -0.34 | 0.27 | -0.59 | 0.05 | -0.25 | 0.43 |
|  |  |  | After | -0.36 | 0.24 | -0.46 | 0.13 | -0.49 | 0.10 |
| Quantitative Imaging |  | VSI_mean | Before | 0.05 | 0.86 | 0.30 | 0.33 | 0.37 | 0.22 |
|  |  |  | After | 0.22 | 0.48 | 0.58 | 0.05 | 0.42 | 0.17 |
|  |  | VSI_cv | Before | -0.01 | 0.95 | -0.10 | 0.74 | 0.04 | 0.89 |
|  |  |  | After | -0.14 | 0.66 | -0.45 | 0.14 | -0.66 | 0.06 |
|  |  | QSI_mean | Before | 0.13 | 0.68 | 0.16 | 0.60 | -0.02 | 0.93 |
|  |  |  | After | 0.67 | **0.01** | 0.30 | 0.34 | 0.50 | 0.09 |
|  |  | QSI_cv | Before | -0.09 | 0.77 | -0.30 | 0.33 | 0.00 | 0.98 |
|  |  |  | After | -0.31 | 0.31 | -0.49 | 0.09 | -0.66 | **0.01** |

Note: data are the rho correlation coefficient of Spearman.
Legends: ABPA= allergic bronchopulmonary aspergillosis; FEV1=forced expiratory volume in 1 second; FVC=forced volume capacity; FEF25-75=forced expiratory flow at 25%-75%; %p=percentage of predicted value; MR= magnetic resonance; V= ventilation; Q= perfusion; VSI= ventilation signal intensity; QSI= perfusion signal intensity; cv= coefficient of variation

**Table E3.**
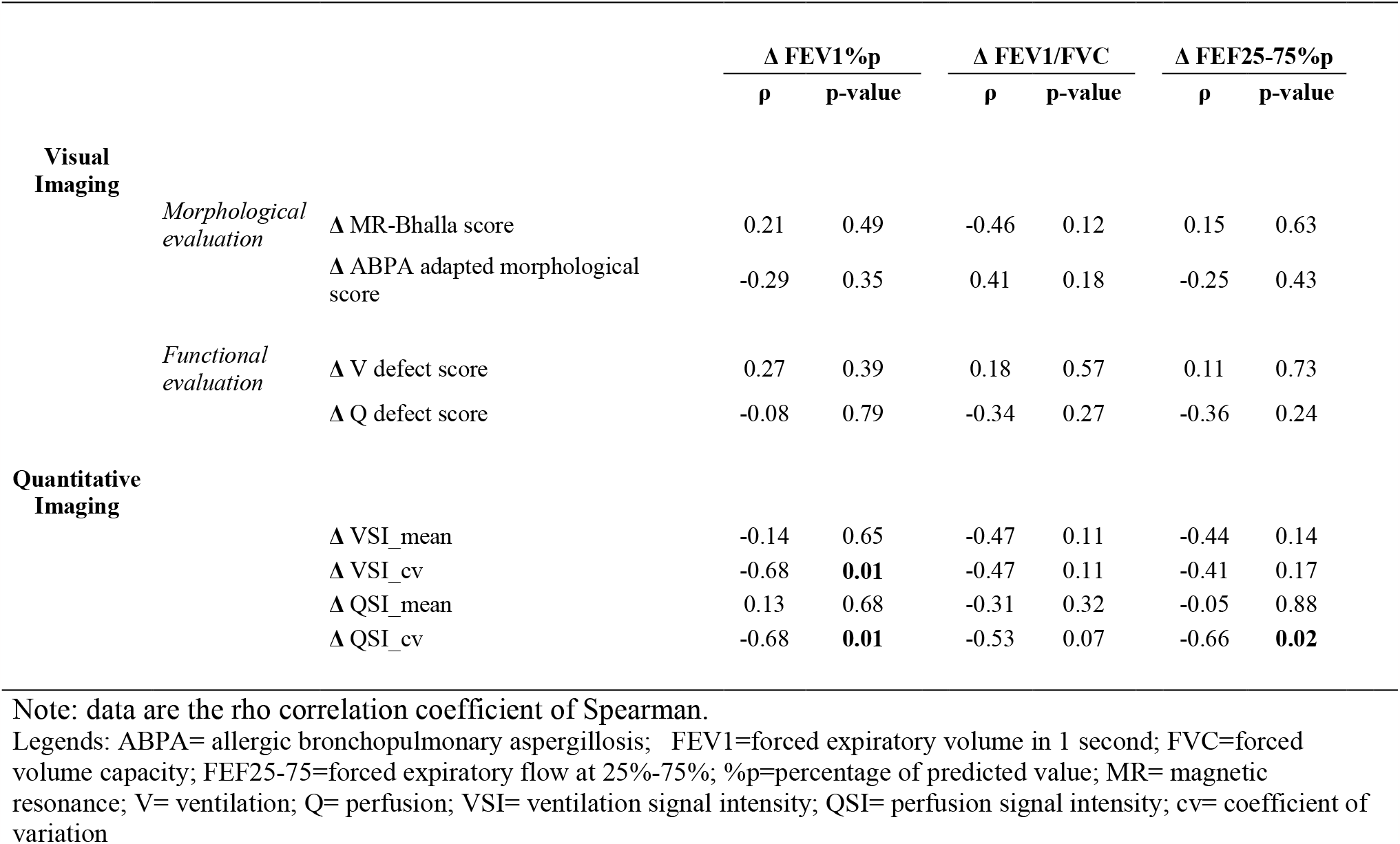
Correlation between the variation of MRI metrics with the variation of pulmonary function test before and after treatment of ABPA.

**Table E4.**
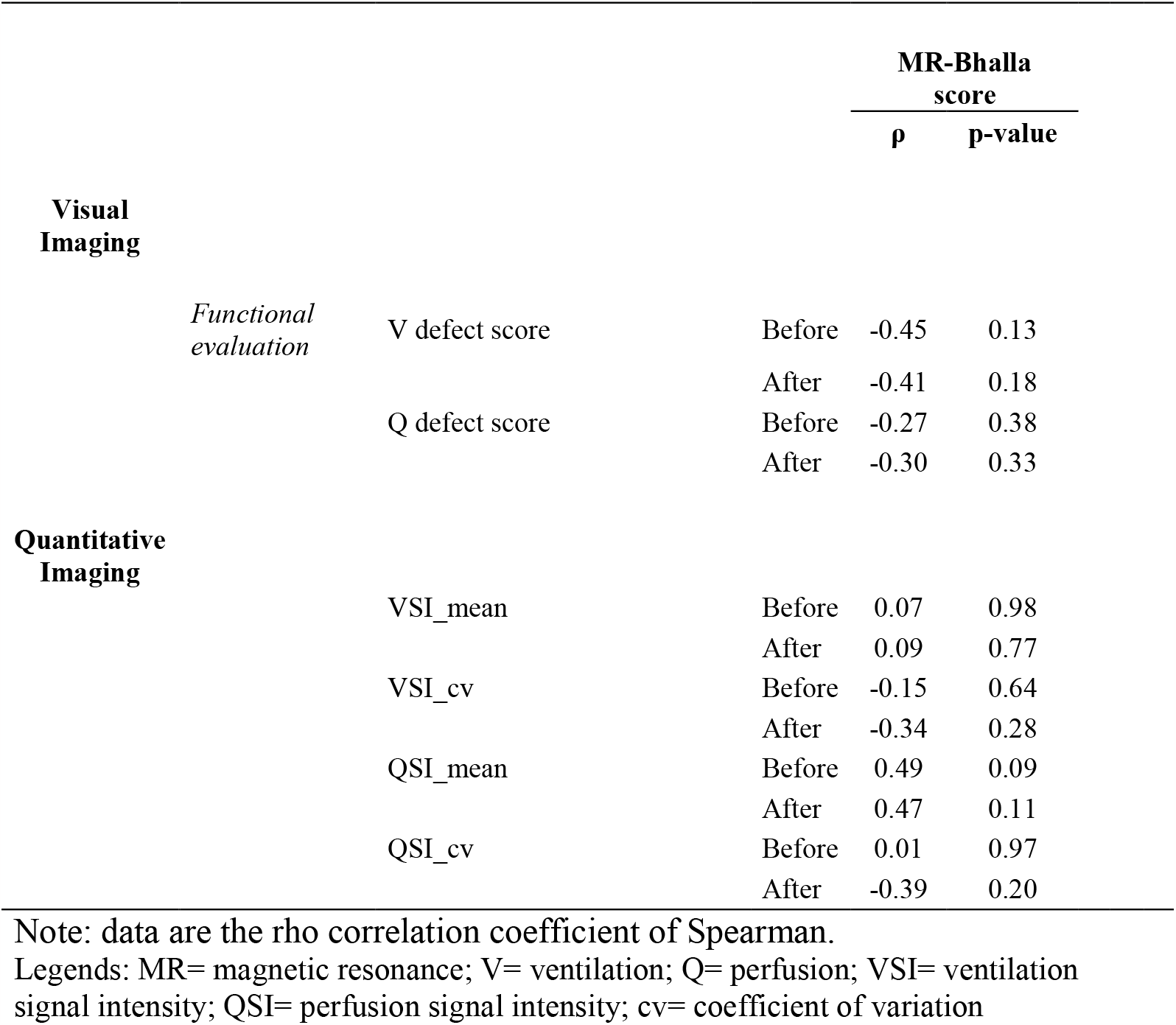
Correlation between MR-Bhalla score and both visual and quantitative FD lung MRI.

## Notes

**Grant support:** Ilyes Benlala received academic funding from la Fondation du Souffle (Lauréat 2021), la Fondation Bordeaux Université-Fonds Délorme Broussin and IdEx Bordeaux LMU-Bordeaux Research Cooperation Program

### Competing Interest Statement

F. Laurent reports personal fees and non-financial support from Chiesi, Boehringer, AstraZeneca, Bayer, Gilead and GSK, outside the submitted work; P. Berger reports personal fees and non-financial support from AstraZeneca, GSK, Novartis, Chiesi and Boehringer, and personal fees and non-financial support from Menarini and Sanofi, outside the submitted work.

### Funding Statement

Ilyes Benlala received academic funding from la Fondation du Souffle (Laureat 2021), la Fondation Bordeaux Universite-Fonds Delorme Broussin and IdEx Bordeaux LMU-Bordeaux Research Cooperation Program

### Author Declarations

Ethics committee/IRB of University Hospital of Bordeaux gave ethical approval for this work

### Summary of Updates

Competing interest statment was updated

